# Identifying compounds to treat opiate use disorder by leveraging multi-omic data integration and multiple drug repurposing databases

**DOI:** 10.1101/2024.01.17.24301329

**Authors:** Jeran K. Stratford, Megan Ulmer Carnes, Caryn Willis, Melyssa S. Minto, Logain Elnimeiry, Ravi Mathur, Matthew Schu, Bryan C. Quach, Javan Carter, Tracy Nolen, Nathan Vandergrift, Thomas Kosten, Eric Otto Johnson, Bradley T. Webb

## Abstract

Genes influencing opioid use disorder (OUD) biology have been identified via genome-wide association studies (GWAS), gene expression, and network analyses. These discoveries provide opportunities to identifying existing compounds targeting these genes for drug repurposing studies. However, systematically integrating discovery results and identifying relevant available pharmacotherapies for OUD repurposing studies is challenging. To address this, we’ve constructed a framework that leverages existing results and drug databases to identify candidate pharmacotherapies.

For this study, two independent OUD related meta-analyses were used including a GWAS and a differential gene expression (DGE) study of post-mortem human brain. Protein-Protein Interaction (PPI) sub-networks enriched for GWAS risk loci were identified via network analyses. Drug databases Pharos, Open Targets, Therapeutic Target Database (TTD), and DrugBank were queried for clinical status and target selectivity. Cross-omic and drug query results were then integrated to identify candidate compounds.

GWAS and DGE analyses revealed 3 and 335 target genes (FDR q<0.05), respectively, while network analysis detected 70 genes in 22 enriched PPI networks. Four selection strategies were implemented, which yielded between 72 and 676 genes with statistically significant support and 110 to 683 drugs targeting these genes, respectively. After filtering out less specific compounds or those targeting well-established psychiatric-related receptors (*OPRM1* and *DRD2*), between 2 and 329 approved drugs remained across the four strategies.

By leveraging multiple lines of biological evidence and resources, we identified many FDA approved drugs that target genes associated with OUD. This approach a) allows high-throughput querying of OUD-related genes, b) detects OUD-related genes and compounds not identified using a single domain or resource, and c) produces a succinct summary of FDA approved compounds eligible for efficient expert review. Identifying larger pools of candidate pharmacotherapies and summarizing the supporting evidence bridges the gap between discovery and drug repurposing studies.

## INTRODUCTION

### Significance

There were more than 80,000 opioid overdose deaths in the United States in 2021 which is 10 times higher than in 1999 and largely due to synthetic opioids^1–3^. The societal and economic burden of the opioid epidemic is immense. Currently, opioid use disorder (OUD) has only three FDA approved drugs for medication assisted treatment (methadone, naltrexone, and buprenorphine^4^) and one additional detoxification agent, lofexidine^5^. The number of off-label drugs commonly used in substance use disorder (SUD) treatment programs are also limited and include baclofen, clonidine, divalproex, gabapentin, ondansetron, topiramate^6^. In contrast, type 2 diabetes has at least 37 different approved pharmacotherapies representing 10 separate classes^7^. As such, there is a pressing need to identify new OUD pharmacologic targets for drug development and quickly translate these findings into medication-assisted therapies. However, psychiatric drug development overall has been slow and outpaced by other areas of medicine^8^. One strategy to address the need for additional OUD pharmacotherapies is to identify drugs approved for other conditions that target biological systems implicated in OUD. Approved drugs have known dosing and safety profiles and can be brought to the market faster and less expensively than novel compounds through drug repurposing studies. In this work, the terms compounds, drugs, and pharmacotherapies are equivalent and do not necessarily reflect development stage or approval status.

### Catalog of ‘failed’ compounds and *in silico* identification

Many compounds pass safety (Phase II) but not efficacy trials (Phase III). A recent study of 21,143 compounds in phase II trials showed success rates of 48.6% for progressing to phase III and only 21% to FDA approval^9^. This large collection of ‘failed’ compounds are likely safe and could be efficacious treatments for other conditions such as OUD and other SUDs for which there is a significant need for new treatments. Compound identification approaches that do not require new data generation have been labelled *in silico*. For this work, we limit this definition to identifying known compounds in data databases linked to genes by leveraging results from existing omic studies for repurposing studies. Empirical evidence supports using these strategies since drugs with targets based on genetic support succeed twice as often in clinical trials^10,11^, and other molecular data (e.g., transcriptomic and proteomic) can inform biological processes impacted by OUD within relevant tissue types. Fortunately, there is a growing array of resources that can be applied to identify these potential OUD treatments.

### Leveraging discovery biology to accelerate treatment

While various lifestyle factors and environmental exposures contribute to risk, OUD also has a genetic component with heritability estimates of 54–60%^12,13^ and some genetic risk is shared with other SUDs and psychiatric disorders^12,14–17^. Recently, specific genetic variation influencing the risk of developing OUD have been identified^18–23^. While genetics offers critical insight into the factors that predispose individuals to OUD, other approaches can inform the biological processes occurring across the course of the disorder. Gene expression studies of postmortem human brains represent an opportunity to get closer to the disease state and elucidate the etiology of OUD and comorbid conditions in ways not possible using genetics alone. There is strong evidence for acute and persistent changes in central nervous system gene expression resulting from exposure to substances of abuse^24,25^, stress^26^, and their associated disorders including OUD^27^, alcohol use disorder (AUD)^28,29^, and post-traumatic stress disorder (PTSD)^30,31^.

Integrating genetic and expression studies can improve identification of relevant genes^32,33^. Both human and animal model gene co-expression networks can be further validated for a role in SUDs through co-analysis^34–36^. Furthermore, statistical approaches have been developed for integrated analysis of gene expression data and GWAS results^32^, and applied to SUD-related data^33^, to identify targets for future therapeutic development^37^.

Network analysis for target identification offers opportunities beyond single target identification. Compounds that lead to efficacious treatment do not necessarily need to directly impact genes associated with liability and/or consequences of the disorder and may act elsewhere in the network. Indeed, most psychiatric risk loci identified to date are not direct targets of effective pharmacotherapies. While molecular networks such as gene co-expression and Protein-Protein Interaction (PPI) networks offer additional insight beyond genetic risk, integrated analyses are still uncommon. Single or integrated multi-domain investigations rarely focus on producing prioritized targets for subsequent translational studies such as drug repurposing trials.

### Drug Development and Repurposing Databases

In parallel to the growing catalog of biological evidence, there are an expanding set of resources that consolidate and summarize disparate information related to pharmacotherapies and their known or potential gene targets. These resources include tools for investigators to readily query genes or compounds of interest. However, the multiple competing resources have overlapping and unique information and learning the strengths of each resource requires an investment of time. While powerful, systematically querying these resources in an agnostic and reproducible way for many genes with robust evidence can be challenging. Looking up a moderate number of genes across resources is feasible but is time consuming and likely not to be reproducible across investigators. As omic investigations of OUD and other complex psychiatric disorders increase in size and power, so do the number of robustly significant associated genes and the challenge to follow them up in translational studies.

### Summary

To address this challenge, we constructed a framework to identify biological targets and candidate pharmacotherapies for preclinical and clinical OUD repurposing studies. Candidate identification is accomplished by 1) leveraging extant results, 2) performing cross-omic network-based analysis, 3) creating an integrated catalogue of results and biological targets, and 4) generating a prioritized list of existing compounds for future repurposing studies.

## METHODS

Biological data processing and integration: Multiple OUD studies using different biological measures were combined to explore evidence of OUD associations with multi-omic support in humans, including 1) gene-level summary statistics generated from meta-analysis of genome-wide association studies (GWAS) of OUD related phenotypes, 2) differential gene expression (DGE) meta-analysis of opioid overdose death (OOD) in human prefrontal cortex (PFC), and 3) Protein-Protein Interaction (PPI) network analyses.

Gene-level summary statistics: For genetic risk loci, we began with GWAS summary statistics from the largest Genomic Structural Equation Modeling (gSEM)^38^ based OUD meta-analysis to date. The Genetics of Opioid Addiction Consortium (GENOA) gSEM based meta-analysis of an OUD common factor model (gSEM) model included 23,367 cases, 384,629 controls, and an effective sample size of 88,114 (due to non-independence) using four primary GWAS^20^. These include the a) non-gSEM component of GENOA^20^, which is a meta-analysis across 15 European ancestry cohorts, b) a previous OUD meta-analysis of Million Veterans Program (MVP), the Study of Addiction: Genetics and 80 Environment (SAGE), and Yale-Penn (YP) (MVP-SAGE-YP)^23^, c) the Psychiatric Genetics Consortium Substance Use Disorder Group (PGG-SUD)^22^, and the Partners Health Group^18^. Gene-level summary statistics were calculated from GWAS summary statistics using MAGMA v1.08^39^ with a 10 kilobase (kb) window and 1000 Genomes Project EUR reference.

Differential gene expression: We also utilized the largest meta-analysis of human post-mortem prefrontal cortex (PFC) differential gene expression (DGE) to date which combined four published RNAseq studies of opioid overdose death (OOD)^40–43^. The DGE meta-analysis included 285 independent decedents, evaluated 20,098 genes, and yielded 335 genes with significant DGE (FDR q<0.05)^44^. Briefly, each RNAseq expression dataset was separately analyzed using a common QC pipeline followed by DGE analysis via the limma-voom regression framework^45–47^ prior to meta-analysis using weighted Fisher’s method to account for variability in sample sizes across studies.

For this current study, a novel cross omic meta-analysis was conducted by combining the DGE and gene-based GWAS *p*-values using Fisher’s method implemented in R package *metapro*^48^. This DGE-GWAS meta-analyses was performed to detect loci that may not be robustly significant in a single domain (DGE or GWAS) but where the combined evidence from independent analyses is robust. False Discovery Rate (FDR) based thresholds were used instead of family*-*wise error rate adjustments methods such as Bonferroni corrections due to the non-independence of statistical tests in genome-wide analyses. Additionally, FDR thresholds are empirically derived from each set of test statistics. FDR based q-values^49^ for each gene were determined using the R package *qvalue* [<u>10.18129/B9.bioc.qvalue</u>].

Protein-Protein Interaction: Finally, PPI network analyses were performed using dense module GWAS (dmGWAS v3.0)^32,50^ with the MAGMA based GWAS gene-level summary statistics and a PPI network from the Integrated Interactions Database for all human tissues^51^. dmGWAS searches across an entire PPI network to identify localized subsets of interactions with GWAS results that are unlikely due to chance. By dmGWAS nomenclature, these sub-networks are called *modules* and deemed *dense* if they demonstrate enrichment of statistical signals, hence *dense module*. Due to dmGWAS exploring a large space of potential modules, two independent meta-analysis GWAS (non gSEM GENOA and MVP12) were analyzed separately. The non-gSEM component of GENOA [PMID: 36207451] is a meta-analysis across 15 European ancestry cohorts (N=304,507, Case N=7,109). MVP12^23^ is a GWAS meta-analysis of parts 1 and 2 of the Million Veteran Program (N=79,719, Case N=8,529). Having two independent dmGWAS results allowed bidirectional discovery and validation analysis using the dmGWAS dualEval function. Significant modules that were replicated in the independent validation dataset via dualEval were collapsed into summary PPI modules. Graphing and visualization of summary modules was performed using the R package *cytoscape* v3.9.1^52^.

### Gene Selection Strategies

To develop a strategy to select and rank genes across domains, we sought to balance the relative contribution of each source with the downstream goal of identifying approved compounds targeting priority genes with robust evidence in both single and across domains. Four strategies were used to assess the impact of varying levels of evidence for association with OUD on the number of genes brought forward for target compound identification (Table 1). Genes were selected based on evidence from four domains including 1) gene-level summary statistics generated from meta-analysis of GWAS of OUD related phenotypes, 2) DGE meta-analysis of OOD in human PFC, 3) a cross DGE-GWAS meta-analysis, and 4) PPI networks enriched for GWAS loci. Within-domain FDR q-values were calculated and were the basis for the four strategies which are detailed in Table 1. FDR q-values are more appropriate than Bonferroni correction for multiple testing in large-scale omic studies where there is extensive non-independence of statistical tests.

**Table 1:** Gene selection strategies and FDR thresholds. GWAS is gene-based MAGMA results.

|  | Inclusion summary |  |  |  |
| --- | --- | --- | --- | --- |
|  | FDR threshold by domain |  |  |  |
| Strategy | DGE | GWAS | DGE-GWAS meta | PPI GWAS network included? |
| Strict, no network | 0.01 | 0.01 | 0.01 | No |
| Strict plus network | 0.01 | 0.01 | 0.01 | Yes |
| Hybrid | 0.01 | 0.16 | 0.01 | Yes |
| Significant | 0.05 | 0.05 | 0.05 | Yes |

The first strategy (Strict, no network) required robust significant evidence (q<0.01) in at least one domain and did not include the network-based genes. The second strategy (Strict plus network) added genes found in enriched PPI network modules by dmGWAS to the first strategy. To balance the contribution of genes from DGE and GWAS, a third strategy (Hybrid) selected genes a) with robust DGE or DGE-GWAS meta-analysis evidence (q<0.01), or b) suggestive GWAS evidence (q<0.16), or c) were in enriched PPI networks. The suggestive threshold of 0.16 is the *p*-value threshold corresponding to Akaike Information Criterion(AIC)^53^ and has previous been used as suggestive threshold in other gene-wise analyses^54^. For the last strategy (Broad), genes were included if they were a) genome-wide significant (GWS) based on an FDR q-value threshold of 0.05 for any domain (DGE, gene-level GWAS, and DGE-GWAS meta-analysis) or b) were in enriched PPI networks. For each strategy, genes were ranked based on the minimum *p*-value across the 3 non-network domains.

### Drug Repurposing Databases and Compound Identification

To comprehensively summarize existing pharmacotherapies targeting known genes irrespective of relationship to OUD, four large drug related resources were queried independently including Pharos a web interface to browse the Target Central Resource Database (TCRD, http://juniper.health.unm.edu/tcrd/)^55,56^, Open Targets^57^, Therapeutic Target Database (TTD)^58^, and DrugBank^59^ to generate comprehensive cross-resource drug summaries including clinical status (approved), target selectivity (total number of targets), and other existing metrics (approved uses, safety). For Pharos and Open Targets, custom queries leveraging existing database specific APIs were written to extract this information, while for TTD and DrugBank downloads were queried. CHEMBL identifiers were used to link information across drug databases. Cross-resource summaries were generated for every protein coding gene (n=22,842) based on GENCODE^60^ V41 with an available Ensembl ID^61^.

### Identification of approved drugs targeting biologically supported genes

Integrated biological evidence and drug database query summaries were merged via Ensembl ID (version 107). For each strategy, genes were considered passing and taken forward if they met at least one OUD related inclusion threshold in a strategy. These passing gene sets were used as the input set to determine drugs which targeted them.

Since a drug can target more than one gene, we sought to assess how specific a drug was to these OUD related genes. If a drug targeted at least one passing gene, the relative specificity of the drug to OUD related genes was defined and calculated as the number of passing genes/total number genes targeted by the drug. Drugs with a relative specificity less than 0.10 were considered non-specific since more than 90% of the gene targets were not considered OUD related as defined by the inclusion strategy.

## RESULTS

### Summary drug database compounds and gene targets

As a first step in identifying candidate pharmacotherapies for OUD repurposing trials, a systematic search across four major drug databases was conducted to extract all known approved drugs and investigational compounds, their development stage, and potential gene targets. The results of searching Pharos^55,56^, Open Targets^57^, TTD^58^, and DrugBank^59^ in June 2022 yielded 38,882 non-unique compound entries across development stages including *Approved*, *Investigative* (clinical trial), *Experimental* (preclinical drugs), and *Withdrawn* (Table 2). Post-query processing and harmonization resulted in 7,014 compounds with unique CHEMBL identifiers and targets a protein coding gene with an Ensembl ID. As shown in Table 2, no single database contained all or most available compounds and drugs. Drug Bank and Open Targets had the most compounds with 3,940 (56.2%) and 4,015 (57.2%), respectively. However, only 691 compounds were found across all four resources as shown in Figure 1.

**Figure 1:**
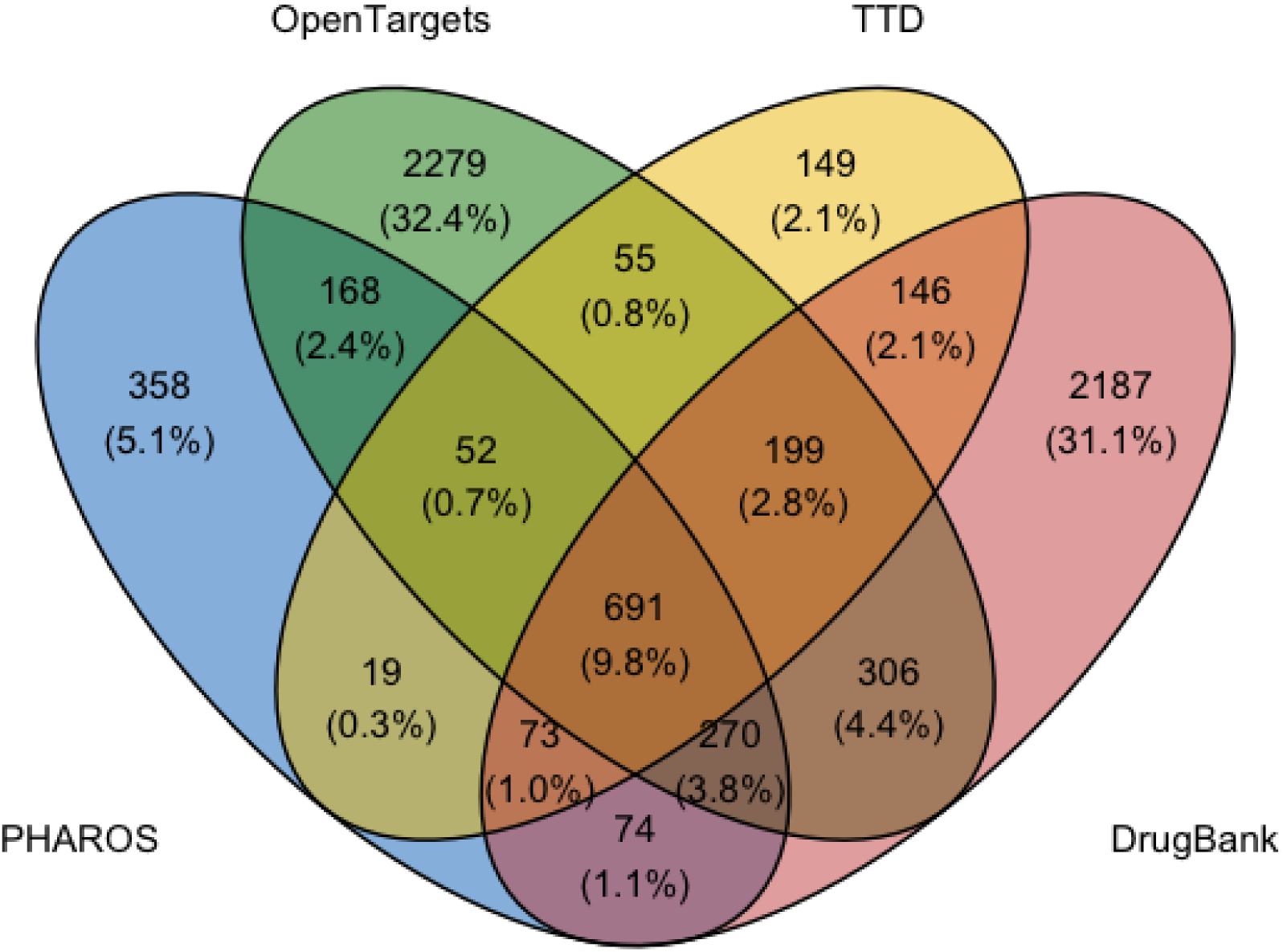
Overlap across drug databases including Pharos (blue n=1,705), Open Targets (green, n=4,020), TTD (yellow, n=1,384) and DrugBank (red, n=3,946).

**Table 2:** Summary of compounds by database by development stage.

| Source | Approved | Investigational | Experimental | Withdrawn | Total | Unique<br>ChEMBL<br>IDs | Has<br>ChEMBL &<br>ENSEMBL ID |
| --- | --- | --- | --- | --- | --- | --- | --- |
| PHAROS | 1,737 | - | - | - | 1,737 | 1705 | 1704 |
| Open Targets | 1,926 | 1,999 | 8 | 87 | 4,020 | 4020 | 4015 |
| TTD | 1,645 | 21,228 | 4,500 |  | 27,373 | 1384 | 1384 |
| Drug Bank | 1,960 | 861 | 2,916 | 15 | 5,752 | 3946 | 3940 |
| Total* | 7,268 | 24,088 | 7,424 | 102 | 38,882 | 7026 | 7014 |

\*Total is sum across sources and does not represent unique compounds.

### Summary of compiled OUD genomic data

We collected two large systematic investigations of OUD biology in humans including GWAS and post-mortem brain (dorsolateral prefrontal cortex) DGE meta-analyses. These results were used to perform network analyses, cross-domain DGE-GWAS meta-analyses, and gene selection by single and multi-domain support. To ensure consistency, we used the MAGMA gene-wise results produced by Gaddis and colleagues as previously reported^20^. False Discovery Rate analysis of these 15,977 gene-wise *p-* values yielded 3 and 115 genes at strict (q<0.05) and liberal (q<0.16) thresholds, respectively (Table 3). Previous studies have demonstrated that genes passing a strict threshold generally explain a fraction of the expected heritability. Using a liberal FDR threshold of 0.16 will on yield approximately 97 true positives at the expenses of including 18 false positives on average.

**Table 3:**
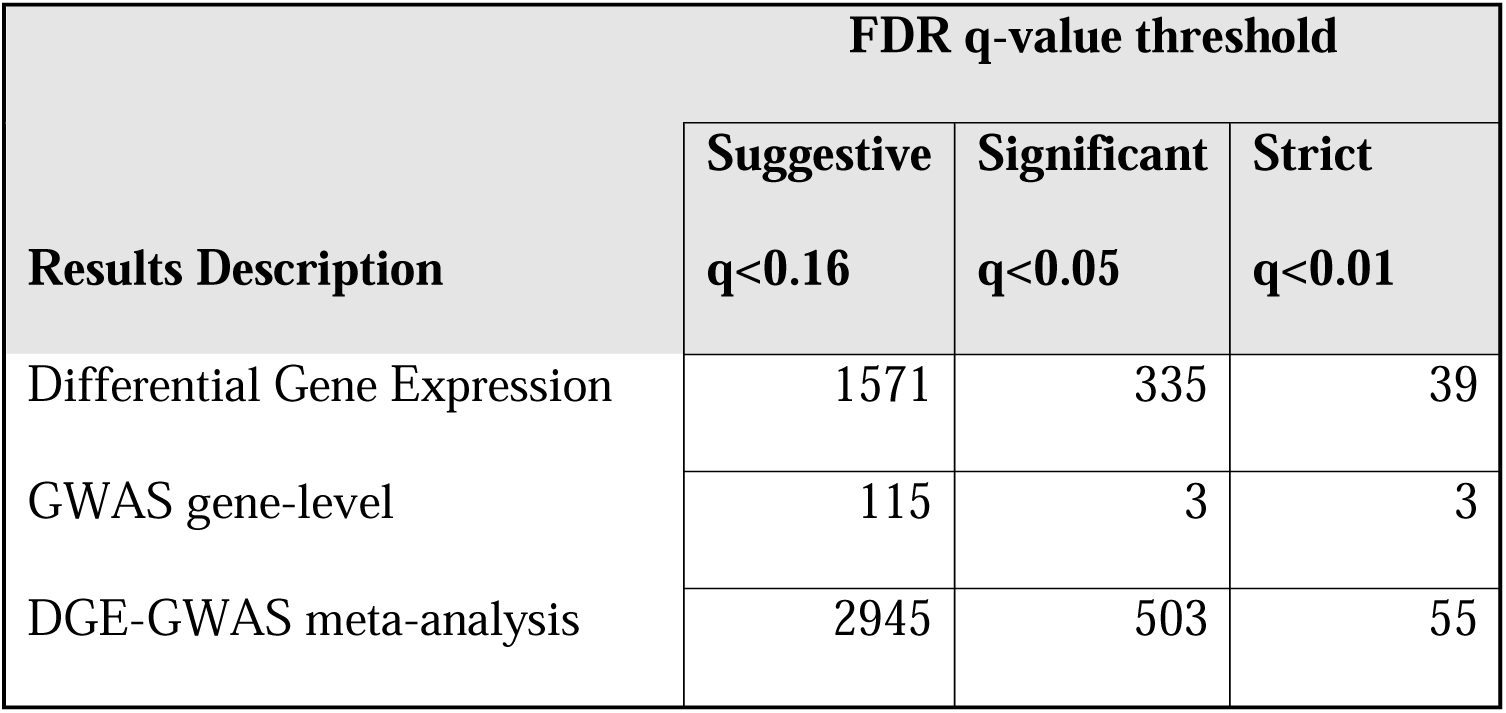
Gene counts by FDR thresholds across evidence sources.

| **Results Description** | **FDR q-value threshold** |  |  |
| --- | --- | --- | --- |
|  | **Suggestive**<br>**q<0.16** | **Significant**<br>**q<0.05** | **Strict**<br>**q<0.01** |
| Differential Gene Expression | 1571 | 335 | 39 |
| GWAS gene-level | 115 | 3 | 3 |
| DGE-GWAS meta-analysis | 2945 | 503 | 55 |

### DGE meta-analysis

As described elsewhere^44^, the meta-analyses of post-mortem human PFC gene expression differences between opioid overdose death (OOD) cases and controls showed 335 genes with robust differences (q <0.05). When the results were limited to 15,377 canonical protein coding genes, the set was reduced to 286 with ∼13% (38/286) being associated with least one approved compound.

### Protein-Protein Interaction (PPI) networks enriched for OUD liability loci

To broaden the set of target genes and drugs, we searched for physically interacting gene clusters harboring genetic variants influencing risk to OUD liability via PPI network analyses. Dense module searching via dmGWAS and two independent subsets of the OUD GWAS meta-analysis described earlier identified 22 candidate PPI modules through bi-directional replication. Six discovery modules from GENOA replicated in MVP12 while 16 modules replicated when reversed. When combined, these enriched subnetworks contained 70 unique genes with 30, 37, and 3 from GENOA, MVP12, or both, respectively. The modules, genes, and gene-specific statistical evidence is in Supplemental Table S1. While the genes within a replicated module may not have robust association evidence individually, the collection of gene-wise *p-*values is significant.

### Gene selection by FDR threshold and by evidence domain

The number of genes with statistical support varied greatly by FDR threshold and dataset. As detailed in Table 3, strict filtering (q<0.01) yielded few genes in GWAS (n=3) while a defensible but less conservative threshold (q<0.16) yielded ∼19% (2945/15,377) of protein coding genes for the DGE-GWAS meta-analysis. These results informed the statistical significance inclusion thresholds used in the four different cross-domain gene selection strategies shown in Table 4. The most restrictive and inclusive strategies produced between 72 and 676 genes for compound searching, respectively. The proportion of statistically supported genes with at least one approved drug varied between 15.6 to 20.5% across selection strategies (Table 4) and were spread across the genome (Figure 2).

**Figure 2:**
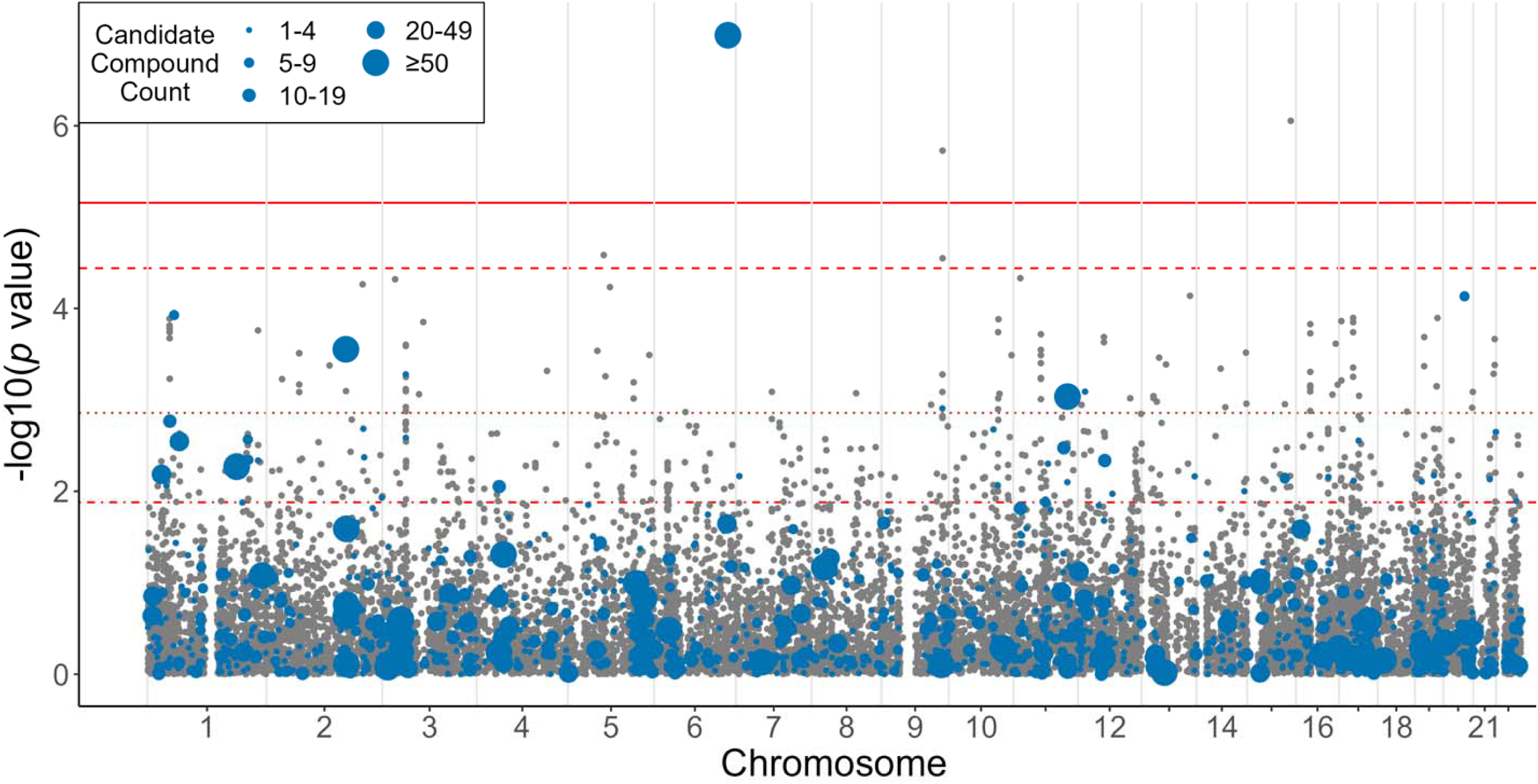
Manhattan plot of gene-level significance. Size of the dot represents the number of drugs that target the gene as listed in the Open Targets drug repurposing database. Red lines are FDR at 5% (solid), 10% (dashed), 20% (dotted) and 40% (dot-dash).

**Table 4:**
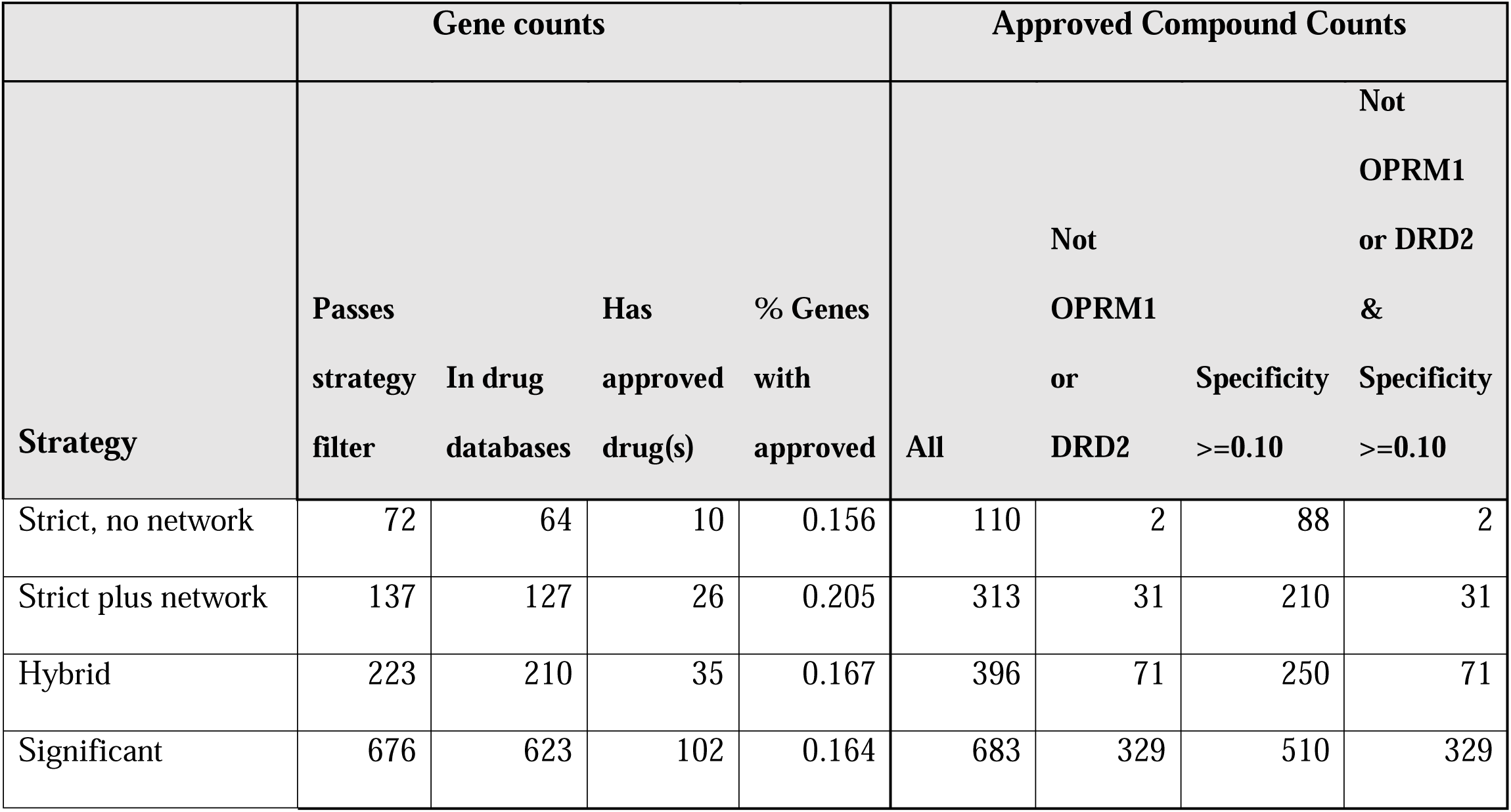
Gene and drug counts by selection strategies.

### Approved compounds with biological support

Ultimately, the four strategies yielded between 110 and 683 compounds targeting empirically supported genes. However, the number of compounds was reduced when excluding non-specific compounds for which empirically supported genes represented <10% of their targets. The full set of identified compounds across strategies with targeted genes and specificity is available in Supplemental Tables S2-6. Inspection of the identified compounds showed that many target *OPRM1* and/or *DRD2*. These genes are the target of extensive drug development and detecting them is evidence that the approach is valid since they may be considered positive controls. To assess if novel or promising repurposing candidates were identified, drugs targeting OPRM1 and DRD2 were filtered out, as summarized in Table 4.

There were thirty-one drugs identified in the “Strict plus network” strategy which did not primarily target *OPRM1* or *DRD2* and were specific to the selected genes. These drugs were inspected and evaluated for plausibility as repurposing candidates for OUD. Most (24 of 31) drugs identified targeted only three genes, *SLCO1B1*, *GUCY1B1*, and *ERBB3*, which each had 11, 10, and 3 approved drugs, respectively. The remaining seven compounds neratinib, bleomycin, cysteamine, tirabrutinib, creatine, esketamine, and entrectinib each targeted a single gene which are *ERBB3*/*MAP4K5*, *LIG1*, *SST*, *TEC*, *CKMT2*, *BDNF*, and *LTK*, respectively.

## DISCUSSION

To facilitate efficient and reproducible searching for candidate OUD pharmacotherapies with biological support from human omic studies for repurposing trials, we implemented a framework for data integration, gene filtering, and systematic searching of four major drug databases. The approach considers evidence from all genes in the human genome and all approved compounds across different resources. This framework is meant to be complementary to the deep information contained within each of the drug databases and may be considered as a generalized framework, capable of investigation of other indications.

### Summary of identified compounds classes

Using this framework, we evaluated four statistically supported strategies, all of which identified compounds with support from at least one biological domain. Broadly, these gene to compound target-pairs can be classified into four categories: *Known, Probable, Novel/unexpected, and Non-specific*. We define *Known* as where there is a large robust collection of supporting evidence and the compound is specifically used for OUD treatment. *Probable* targets-pairs are those where the existing evidence outside that used in this study readily supports the biological plausibility. *Novel/unexpected* target-pairs are those without such obvious support. Finally, the *Non-specific* category may include gene-compound pairs where the specificity for OUD-associated genes is low. There may be other gene-compound pairings that don’t fit into these categories including weak evidence linking a gene and drug pair, database error due to non-unique gene symbols used in scientific literature, non-specificity, or specific evidence showing lack of efficacy. While this is a non-exhaustive list of reasons and subjective, it is useful to evaluate the relative success and usefulness of the approach.

### Known compounds

We note that the primary identification framework was designed and implemented without regard for the *Known* category of pharmacotherapies. Still, both *OPRM1* and *DRD2* were identified as having converging evidence. *OPRM1* is both a primary target of opioids and a target for first line treatments for OUD. While there are many pharmacotherapies approved for treating psychiatric disorders that target *DRD2*, they are not considered first-line treatments for OUD. However, there is a known and complex relationship between dopaminergic and opioid neurocircuitry^62^ including dopaminergic D2 receptors being involved in the rewarding properties of opioids^63^ and dopamine receptor antagonists reducing stress induced opiate relapse in rodents^64^. More recently, atypical antipsychotics which are partial agonists of *DRD2* such as brexpiprazole have shown potential as OUD treatments including showing evidence of modulating dopamine-dependent behaviors during opioid use in mice^65^.

### Probable/Expected

In addition to *DRD2* and *OPRM1*, at least one other promising target with an approved compound was revealed. *BDNF* showed robust differential gene expression and remained highly ranked after DGE-GWAS meta-analysis. While Pharos, TTD, and Open Targets did not reveal any approved compounds targeting *BDNF* directly, DrugBank listed esketamine as targeting BDNF. Although esketamine, which is the S form of ketamine, primarily blocks NMDA receptors, this action secondarily modulates *BDNF* through a downstream signaling cascade^66^. Although not listed as target in Drug Bank or Open Targets, both chiral forms of ketamine bind to mu, delta, and kappa opioid receptors^67^. Additionally, giving naltrexone, an opioid antagonist, before giving ketamine will block ketamine’s ability to rapidly reverse depression^68^. Esketamine is being investigated to treat a wide range of psychiatric disorders including major depression, PTSD, SUDs, and opioid withdrawal^69^. The example of BDNF and esketamine shows the need to both search multiple drug resources and follow up large scale automated candidate compound identification to understand why candidate compounds were found.

### Novel/Unexpected

We also identified gene targets and associated compounds that we classify as novel or unexpected. A few examples include neratinib, bleomycin, cysteamine, tirabrutinib, creatine, entrectinib, and the 24 compounds targeting genes *SLCO1B1*, *GUCY1B1*, and *ERBB3*. Neratinib, bleomycin, and entrectinib are all anti-cancer medications. Neratinib primary targets are *MAP4K5* and *ERBB3* as well as 12 non-supported genes (specificity 14.3%) while bleomycin is a cytotoxic chemotherapy targeting DNA ligase I (*LIG1*). Tirabrutinib is a Bruton’s tyrosine kinase (*BTK*) inhibitor approved to treat autoimmune disorders and hematological malignancies^70^ and also targets *TEC* (tec protein tyrosine kinase), which has support from DGE (q=0.077), DGE-GWAS meta-analysis (q=0.043), and network analyses. Entrectinib targets 10 genes including leukocyte tyrosine kinase (*LTK*) which is implicated primarily by network analysis with support from DGE-GWAS meta-analysis (q=0.08). Literature searching did not reveal an existing relationship between the neratinib, bleomycin, tirabrutinib, or entrectinib and opioid receptors or psychiatric outcomes more broadly. However, there is an existing literature exploring the relationship between the mu opioid receptors and cancer^71^.

For *SLCO1B1*, *GUCY1B1*, and *ERBB3*, post-hoc inspection revealed supporting evidence coming primarily from the network analysis. *SLCO1B1* is primary expressed in liver according to GTEx^72^ and was not tested in the DGE likely due to low expression in brain. Given the lack of expression in brain and other supporting evidence, compounds targeting *SLCO1B1* are probably not good candidates for repurposing studies. In contrast, *GUCY1B1* encodes guanylate cyclase 1 soluble subunit beta 1 has widespread expression including brain and is involved with nitric oxide signaling which modulates the actions of opioid related analgesics^73^. According to the Protein Atlas^74^ (proteinatlas.org) expression clustering, *GUCY1B1* is highly correlated with psychiatric genes of interest including *GABRB1* and *HTR2A*. Since the supporting evidence is limited to the human network results, additional evidence from model organisms or other omics would increase confidence that *GUCY1B1* targeting compounds are good candidates from repurposing trials.

*ERBB3* encodes the Erb-b2 receptor tyrosine kinase 3, which is expressed across many tissues including brain, and binds neuregulin 1 (*NRG1*). While *NRG1* has been the subject of many psychiatric genetic investigations and variants in *ERBB3* are associated with smoking initiation^75^, there is no obvious evidence linking *ERRB3* to OUD.

### Nonspecific

While compounds that target many gene products, directly or indirectly, may indeed be good pharmacotherapies, evaluating these is challenging. Given the large catalog of other potential compounds for OUD repurposing trials, we subjectively conclude these are lower priority to investigate. We highlight fostamatinib as one example which targets six supported genes including *DCLK1*, *DYRK1A*, *LTK*, *MAP4K5*, *TAOK2*, and *TEC*. Individually only *MAP4K5* was robustly significant when considering DGE (p=1.92E-05, q=0.0099) or GWAS alone. The other 5 genes were implicated via network analysis and cross DGE-GWAS meta-analysis. While this demonstrates the need to search for converging evidence, fostamatinib was considered a non-specific compound since the cross-database search showed it targeted 299 genes, yielding a specificity of 2%.

### Strength and limitations

To identify established and novel therapeutic targets for OUD, the current study first identified genes with evidence across three domains, which is a strength of this study. The inclusion of a broader array of evidence such as epigenetics, more tissues, and model organisms would likely increase the number of genes targets and thereby the number of candidate drugs for OUD repurposing studies. The strategy implemented chose a focused set of data with the goal of balancing robustness and interpretability. One goal was to allow subject domain experts to evaluate the supporting evidence.

The study also used gene-wise *p-*values which aggregates evidence for many SNPs across a gene into a single statistic. This is necessary to integrate results at the gene level to allow dataset harmonization. While MAGMA accounts for non-independence of individual SNPs, associated SNPs and the aggregate statistic may not represent a true association between the trait and the gene which may be nearby.

Another limitation is the imbalance in evidence across biological domains and the challenge in balancing the contribution of each source. Postmortem gene expression included hundreds of statistically significant loci while GWAS for liability risk loci showed a limited number of genome-wide significant loci. However, there is compelling evidence from heritability and enrichment analyses that the available GWAS results are a mixture distribution of real and null effects. As such we employed a statistically sound FDR based strategy and performed a cross DGE and GWAS meta-analysis. Limiting GWAS evidence to only genome-wide significant loci creates different limitations, and the multidomain strategy used here is unlikely to bring forward loci with suggestive evidence in only a single domain. Importantly, the approach allows prioritization of genes with robust evidence from DGE which represent a complex mix of liability, exposure, neuronal remodeling, severity, and disorder progression.

### Conclusions

Selecting gene targets with the goal of identify promising pharmacotherapies likely benefits from a multi-faceted approach that integrates varied domains of biological evidence and is more fruitful than targeting compounds identified from a single domain. In addition to leveraging multiple biological domains, we demonstrate that querying multiple drug databases provides greater coverage of available candidate pharmacotherapies. This framework of integrating supporting biological evidence and readily summarizing available approved compounds helps to bridge the gap between discovery and translational studies. Ultimately, this addresses the pressing need to identify new targets for OUD drug development and repurposing studies.

## Data Availability

All data produced in the present study are available upon reasonable request to the authors.

## Acknowledgements

The U.S. Army Medical Research Acquisition Activity, 820 Chandler Street, Fort Detrick MD 21702-5014 is the awarding and administering acquisition office. This work was supported by the Office of the Assistant Secretary of Defense for Health Affairs through the Alcohol and Substance Use Research Program under Award No. W81XWH1820044 (PASA 2). Opinions, interpretations, conclusions and recommendations are those of the author and are not necessarily endorsed by the Department of Defense.

